# Time is Brain: Detection of Nonconvulsive Seizures and Status Epilepticus During Acute Stroke Evaluation Using Point-of-Care Electroencephalography

**DOI:** 10.1101/2024.06.24.24309423

**Authors:** Kapil Gururangan, Richard Kozak, Parshaw J. Dorriz

## Abstract

**Objectives:** Seizures are both a common mimic and a potential complication of acute stroke. Although EEG can be helpful to evaluate this differential diagnosis, conventional EEG infrastructure is resource-intensive and unable to provide timely monitoring to match the emergent context of a stroke code. We aimed to evaluate the real-world use and utility of a point-of-care EEG device as an adjunct to acute stroke evaluation.

**Materials and Methods:** We performed a retrospective observational cohort study at a tertiary care community teaching hospital by identifying patients who underwent point-of-care EEG monitoring using Rapid Response EEG system (Ceribell Inc., Sunnyvale, CA) during stroke code evaluation of acute neurological deficits during the study period from January 1, 2020 to December 31, 2020. We assessed the frequency of seizures and highly epileptiform patterns among patients with either confirmed strokes or stroke mimics.

**Results:** Point-of-care EEG monitoring was used in the wake of a stroke code in 70 patients. Of these, neuroimaging and clinical information resulted in a diagnosis of stroke in 38 patients (28 ischemic, 6 hemorrhagic, 4 transient ischemic attack; median NIHSS score of 6.5 [IQR 2.0-12.0]) and absence of any stroke in 32 patients. Point-of-care EEG detected seizures and highly epileptiform patterns in 6 (15.8%) stroke patients and 11 (34.4%) stroke-mimic patients, including 2 patients with persistent expressive aphasia due to repeated focal seizures.

**Conclusions:** Point-of-care EEG has utility for detecting nonconvulsive seizures in patients undergoing acute stroke evaluations.

## INTRODUCTION

Epileptic seizures and acute stroke are commonly encountered entities in acute care settings, and they are frequently among the differential diagnosis of acute neurological deficits.^1,2^ Seizures are common mimics of cerebrovascular accidents, accounting for 4-13% of stroke mimics, and both acute symptomatic seizures and other highly epileptiform patterns can also occur during the hyperacute phase of both ischemic and hemorrhagic strokes.^3–9^ As a result, stroke codes may be activated for stroke-like symptoms caused by seizures (e.g., hemiparesis seen during either ictal or post-ictal state, isolated expressive aphasia caused by focal nonconvulsive status epilepticus, sudden altered mental status due to nonconvulsive seizures in critical illness), transient ischemic attacks (TIAs, including seizure-like limb- shaking TIAs due to internal carotid artery occlusion), or acute strokes with concomitant seizures.^10–14^ In addition, older adults with new-onset seizures may be found on imaging to have a cerebrovascular accident but still require urgent treatment of these acute symptomatic seizures.^15–17^

Electroencephalography (EEG) has been shown to reliably detect both epileptic activity and cerebral injury with high temporal resolution, as well as provide acute and subacute prognostic information in stroke patients.^18–21^ It remains the mainstay for diagnosing nonconvulsive seizures to guide emergent treatment, informing differential diagnosis, detecting other ictal-interictal continuum patterns that may help stratify patients’ risk for post-stroke seizures, and continuously monitoring background activity for post-stroke prognostication.^18,22^ Although it predated computed tomography (CT) and magnetic resonance imaging (MRI) technologies by roughly 50 years, EEG has been largely supplanted by neuroimaging workflows in acute stroke evaluation, in part because conventional EEG monitoring requires trained technologists and bulky machines for acquisition and specialized neurologists for accurate review, all of which are difficult to marshal at the bedside during a stroke code and at all hours.^23,24^ Because of the limited access to EEG in many hospitals, prior studies have investigated the diagnostic utility of various neuroimaging modalities for differentiating seizure and stroke, albeit with mixed results. Both CT perfusion and MRI can show distinct patterns (e.g., focal hyperperfusion and restricted diffusion, respectively, crossing vascular territories or involving subcortical structures such as the thalamus or splenium) specific to ongoing status epilepticus, but these findings are not consistently present depending on the timing of imaging relative to the termination of seizure activity and transition to postictal pseudonormalization and then hypoperfusion.^25–34^ Therefore, there remains a need for rapid EEG devices that can be easily deployed in the acute stroke setting and used in combination with neuroimaging data by the bedside team to detect seizures as stroke mimics with high sensitivity and evaluate for salient electrographic abnormalities in patients with confirmed strokes.

One such rapid EEG device using a point-of-care reduced electrode array was developed by Ceribell Inc., and it now serves as standard-of-care for evaluating patients suspected to have nonconvulsive seizures at many academic and community hospitals.^35^ In this study, we aimed to report on the use of this device as an adjunct to acute stroke evaluation at our center and describe the point-of-care EEG findings in patients diagnosed with either imaging-confirmed strokes or stroke mimics.

## METHODS

### Study Design

We conducted a retrospective observational cohort study of adult patients (age ≥18 years) who underwent at least one episode of point-of-care EEG monitoring with Ceribell’s Rapid Response EEG system at Providence Mission Hospital Mission Viejo, a tertiary care community teaching hospital, between January 1, 2020 and December 31, 2020.^36^ Of 319 patients in our cohort, we identified a subset of 70 patients who underwent point-of-care EEG monitoring as part of standard-of-care during or closely following a stroke code activated due to concern for acute stroke (**Figure 1**). Our local conventional and rapid EEG infrastructure and workflow, as well as our study protocol, were described previously.^36^ The indications for point-of-care EEG monitoring at our hospital consisted of (1) preceding clinical event concerning for seizures without return to neurological baseline despite a reasonable observation period or (2) unexplained encephalopathy; another standard indication for post-resuscitation monitoring was not relevant to this patient subgroup.

**Figure 1.**
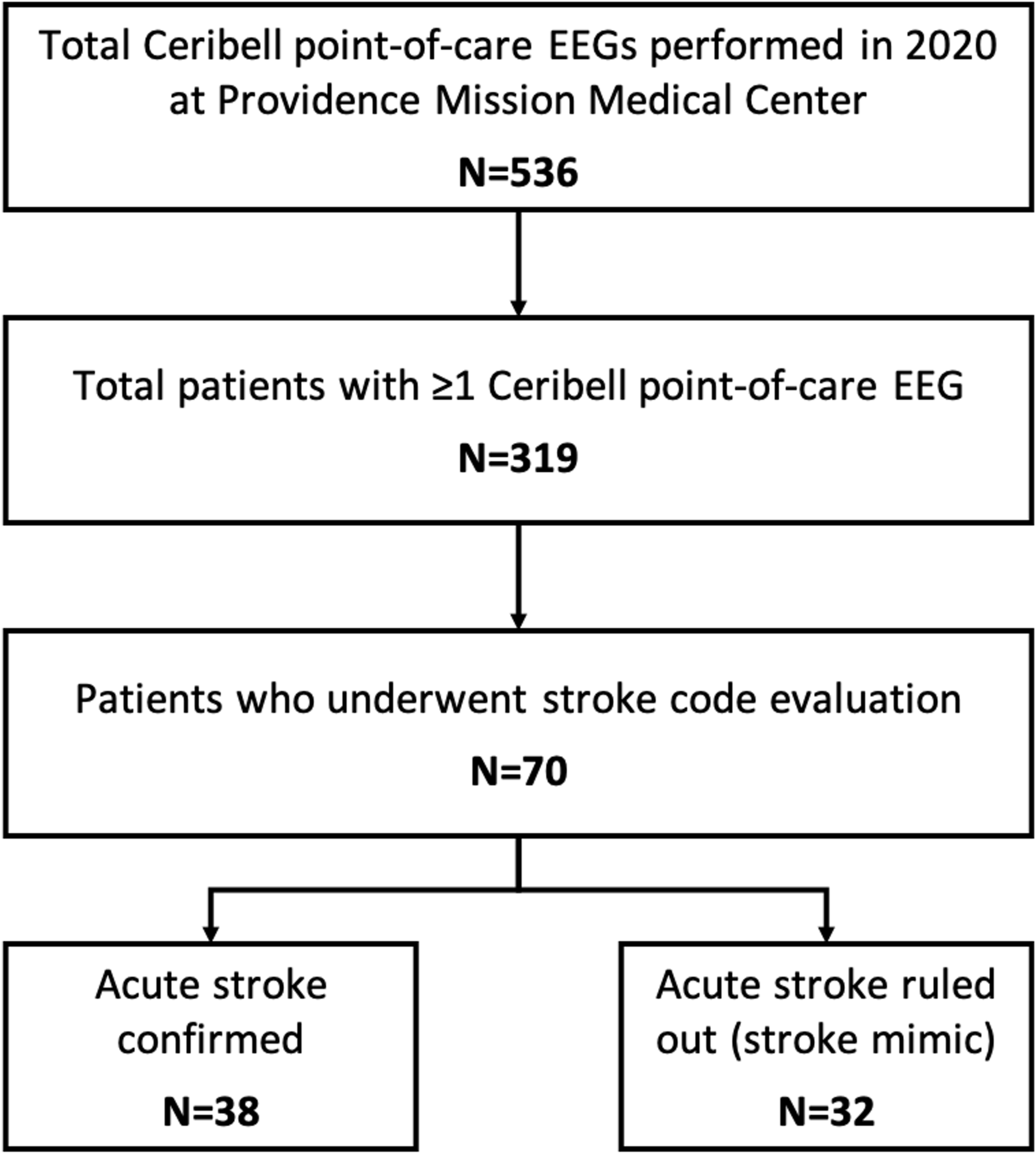
Cohort Selection Flow Diagram.

The study was approved by the Providence Mission Hospital institutional review board (STUDY2021000480) and conducted in accordance with the World Medical Association’s Declaration of Helsinki and Strengthening the Reporting of Observational Studies in Epidemiology (STROBE) guideline.

### Measurements

We performed a chart review of patients’ medical records to obtain demographic and clinical information (e.g., age, sex, initial Glasgow Coma Scale [GCS] score, neurological history, presenting stroke-like symptom, initial National Institutes of Health Stroke Scale [NIHSS] and modified Rankin scale [mRS] scores, presence of an emergent large vessel occlusion [ELVO], and antiseizure medication [ASM] or reperfusion treatments). Because this was a subgroup analysis of a previously studied cohort, certain data (such as full imaging reports) were no longer accessible for review for all patients. However clinical information and critical imaging results described in initial history and discharge diagnoses were reported if available.

Each patient’s final diagnosis (i.e., stroke code outcome) was obtained from the medical record and categorized as either confirmed stroke (including ischemic and hemorrhagic strokes and TIAs) or stroke mimic (including seizures, toxic-metabolic encephalopathy, transient global amnesia, neoplasm, and shock). Particular vascular etiologies (e.g., hypertensive encephalopathy, subdural hematomas) were classified as stroke mimics if there was no evidence of parenchymal infarction/hemorrhage based on clinical documentation. Patients may have been given a final clinical diagnosis of seizures in spite of a non-ictal EEG study.

Consistent with prior studies of the Ceribell EEG system^35,36^, point-of-care EEG findings described in the original study report were categorized as either: seizure or status epilepticus, highly epileptiform patterns (HEP, including epileptiform discharges as well as rhythmic or periodic patterns described in the American Clinical Neurophysiology Society’s 2021 Standardized Critical Care EEG Terminology^37^), slow activity (including generalized rhythmic delta activity and non-epileptiform burst suppression), or normal activity. EEG and imaging findings were categorized based on the clinical documentation and study reports that guided patients’ clinical care; we did not independently review or reinterpret these studies.

### Outcomes

We tabulated the frequency of point-of-care EEG findings, with particular attention to ictal and epileptiform abnormalities, among patients with confirmed strokes and stroke mimics. We also identified exemplar cases of seizures or status epilepticus to describe these clinical vignettes in greater detail.

### Statistical Analysis

Descriptive statistics were calculated (mean [SD] or median [IQR] for continuous variables, number and percentage for categorical variables). Significance testing was not performed due to the descriptive nature of this subgroup report.

## RESULTS

Among the 70 patients who underwent point-of-care EEG monitoring in the wake of acute stroke evaluation, 38 (54.3%) were diagnosed with strokes while 32 (45.7%) were diagnosed with stroke mimics. Characteristics of stroke and stroke mimic patients are described in **Table 1** and **Table 2**, respectively. Mean age for the stroke code cohort was 75.0 (SD 10.9), and 28 (40.0%) were female. The indication for point-of-care EEG monitoring was a preceding seizure-like clinical event in 11 patients (15.7%) and unexplained encephalopathy in 59 (84.3%). Unsurprisingly, a history of cerebrovascular disease was more common among stroke patients (94.7% vs. 68.8%), whereas a history of seizures and a preceding seizure-like clinical event were more frequently documented among patients diagnosed with stroke mimics (31.3% and 28.1% vs. 5.3% and 5.3%, respectively).

**Table 1.** Characteristics of Stroke Patients. Abbreviations: ASM, antiseizure medication; EEG, electroencephalography; ELVO, emergent large vessel occlusion; GCS, Glasgow Coma Scale; NIHSS, National Institutes of Health Stroke Scale; TIA, transient ischemic attack.^a^Presenting stroke-like symptoms (as documented in initial history or discharge diagnoses) were categorized as altered mental status, focal weakness (including hemiparesis and isolated facial droop or unilateral limb weakness), focal numbness (including hemisensory loss and unilateral limb numbness), aphasia, dysarthria, gaze preference, or convulsions. If a patient presented with multiple symptoms (e.g., altered mental status and convulsions, or aphasia and focal weakness), then all were included. ^b^Percentage reported as proportion of patients diagnosed with ischemic stroke.

|  | **Stroke (n=38)** |
| --- | --- |
| **Age (years), mean (SD)** | 75.2 (9.2) |
| **Female sex, n (%)** | 17 (44.7) |
| **GCS score, median (IQR)** | 15.0 (13.0-15.0) |
| **Neurological history, n (%)** |  |
| Seizures or ASM use | 2 (5.3) |
| Brain tumor | 0 (0) |
| Cerebrovascular disease or intracranial hemorrhage | 36 (94.7) |
| Head trauma | 0 (0) |
| **Presenting symptom, n (%)<sup>a</sup>** |  |
| Altered mental status | 11 (28.9) |
| Focal weakness | 19 (50.0) |
| Focal numbness | 2 (5.3) |
| Aphasia | 11 (28.9) |
| Dysarthria | 2 (5.3) |
| Gaze preference | 2 (5.3) |
| Convulsions | 2 (5.3) |
| Not specified | 0 (0) |
| **Indication for point-of-care EEG monitoring, n (%)** |  |
| Clinical event concerning for seizure | 2 (5.3) |
| Unexplained encephalopathy | 36 (94.7) |
| **NIHSS score, median (IQR)** | 6.5 (2.0-12.0) |
| **Stroke type, n (%)** |  |
| Ischemic stroke | 28 (73.7) |
| Hemorrhagic stroke | 6 (15.8) |
| TIA | 4 (10.5) |
| **ELVO, n (%)<sup>b</sup>** | 7 (25.0) |
| **Acute reperfusion treatment, n (%)<sup>b</sup>** |  |
| None | 16 (57.2) |
| Thrombolysis only | 8 (28.6) |
| Thrombectomy only | 2 (7.1) |
| Thrombolysis and thrombectomy | 2 (7.1) |

**Table 2.** Characteristics of Stroke Mimic Patients. Abbreviations: ASM, antiseizure medication; EEG, electroencephalography; GCS, Glasgow Coma Scale; TGA, transient global amnesia. ^a^Presenting stroke-like symptoms (as documented in initial history or discharge diagnoses) were categorized as altered mental status, focal weakness (including hemiparesis and isolated facial droop or unilateral limb weakness), focal numbness (including hemisensory loss and unilateral limb numbness), aphasia, dysarthria, gaze preference, or convulsions. If a patient presented with multiple symptoms (e.g., altered mental status and convulsions, or aphasia and focal weakness), then all were included.

|  | **Stroke mimic (n=32)** |
| --- | --- |
| **Age (years), mean (SD)** | 74.7 (12.8) |
| **Female sex, n (%)** | 11 (34.4) |
| **GCS score, median (IQR)** | 13.0 (6.0-14.3) |
| **Neurological history, n (%)** |  |
| Seizures or ASM use | 10 (31.3) |
| Brain tumor | 1 (3.1) |
| Cerebrovascular disease or intracranial hemorrhage | 22 (68.8) |
| Head trauma | 1 (3.1) |
| **Presenting symptom, n (%)<sup>a</sup>** |  |
| Altered mental status | 17 (53.1) |
| Focal weakness | 11 (34.4) |
| Focal numbness | 0 (0) |
| Aphasia | 6 (18.8) |
| Dysarthria | 0 (0) |
| Gaze preference | 0 (0) |
| Convulsions | 5 (15.6) |
| Not specified | 1 (3.1) |
| **Indication for point-of-care EEG monitoring, n (%)** |  |
| Clinical event concerning for seizure | 9 (28.1) |
| Unexplained encephalopathy | 23 (71.9) |
| **Final diagnosis, n (%)** |  |
| Seizure | 15 (46.9) |
| Toxic-metabolic encephalopathy | 9 (28.1) |
| Hypertensive encephalopathy | 4 (12.5) |
| Subdural hematoma | 1 (3.1) |
| Metastatic cancer | 1 (3.1) |
| TGA | 1 (3.1) |
| Shock | 1 (3.1) |

Stroke type was ischemic in 28 patients (73.7%), hemorrhagic in 6 (15.8%), and TIA in 4 (10.5%). Median NIHSS score was 6.5 (IQR 2.0-12.0), and ELVO was detected in 7 patients (25.0% of ischemic strokes).

ELVO patients had higher median NIHSS scores (8.0, IQR 7.0-12.0) compared to non-ELVO patients (5.0, IQR 2.0-13.0). Among patients with acute ischemic stroke, 12 patients (42.8%) received acute reperfusion therapies (thrombolysis alone in 8, thrombectomy alone in 2, and both thrombolysis and thrombectomy in 2). The 16 patients (57.2%) who did not receive acute stroke treatments were excluded from these treatments due to either symptomatic improvement (n=5), last known well time outside of treatment time windows (n=3), anticoagulant use (n=2), recent intracranial hemorrhage (n=1), thrombocytopenia (n=1), recent major surgery (n=1), or patient refusal of treatment (n=1); exclusion criteria were unspecified for two patients. No patients were excluded from acute stroke treatment due to concomitant seizures. Hemorrhagic conversion occurred in two patients with ischemic strokes, one of whom received thrombolysis. One patient treated with thrombolysis underwent decompressive hemicraniectomy. Seizure was the most common stroke mimic, occurring in 15 (46.9%) patients, followed by toxic-metabolic (n=9, 28.1%) and hypertensive (n=4, 12.5%) encephalopathies. Two patients received thrombolysis who subsequently were diagnosed with a stroke mimic.

Point-of-care EEG findings for stroke and stroke mimic patients are shown in **Figure 2**. Seizures or HEP were detected in 6 stroke patients (15.8%), including 2 found to be in electrographic status epilepticus, and 11 stroke mimic patients (34.4%), 2 of whom were found to have focal electroclinical seizures characterized by expressive aphasia. Clinical vignettes of these 4 ictal cases are summarized in **Figure 3**. Among the 15 patients diagnosed with seizure as a stroke mimic (based on all available clinical information, not EEG data alone), point-of-care EEG confirmed seizures in 2 and revealed HEP in 3 (2 with generalized periodic discharges, 1 with lateralized periodic discharges), slowing in 9 (5 focal, 4 generalized), and normal activity in 1.

**Figure 2.**
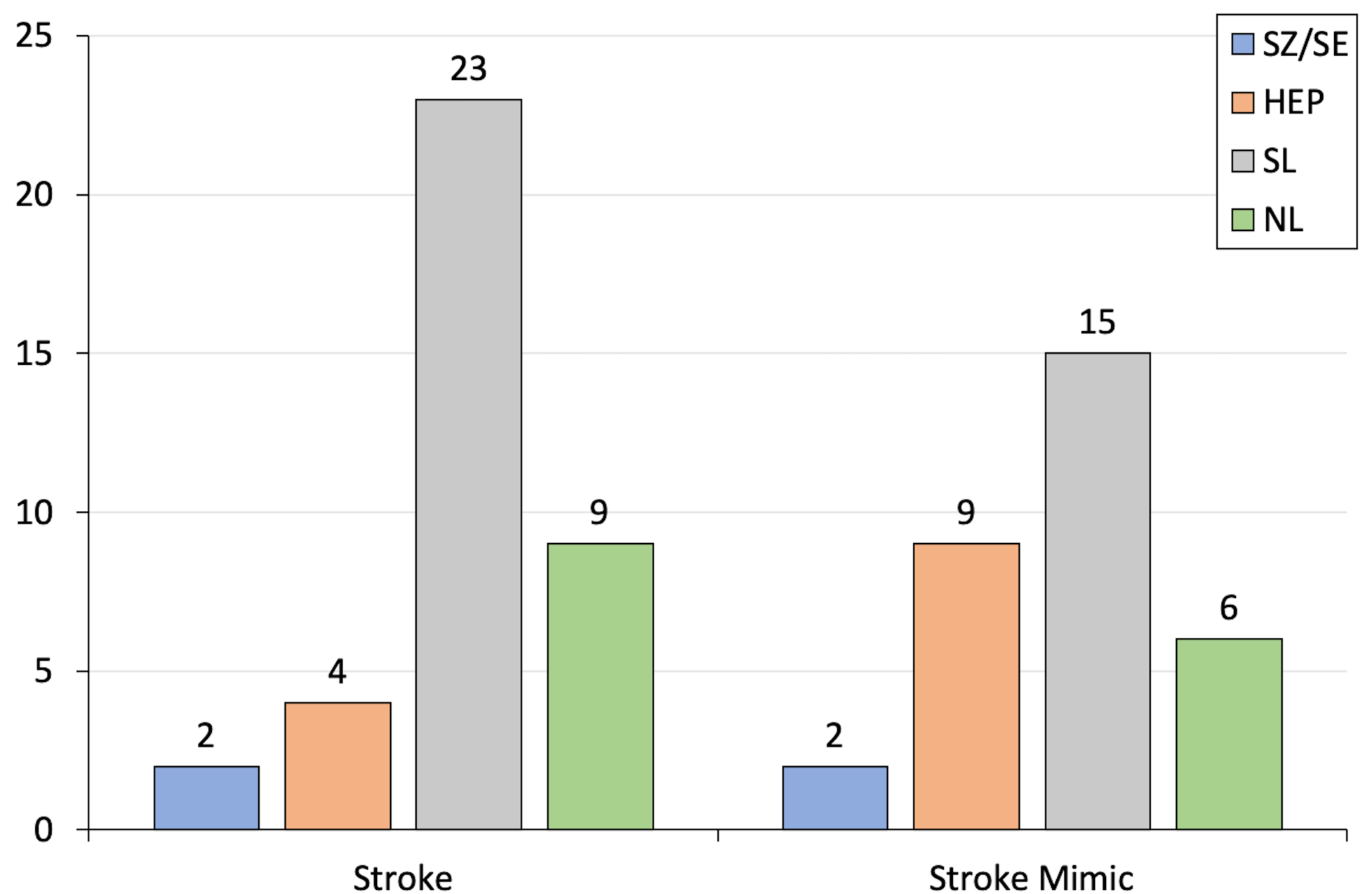
Point-of-Care EEG Findings During Stroke Codes. Abbreviations: EEG, electroencephalography; HEP, highly epileptiform pattern; NL, normal activity; SL, slow activity, polymorphic diffuse or focal; SZ/SE, seizure or status epilepticus.

**Figure 3.**
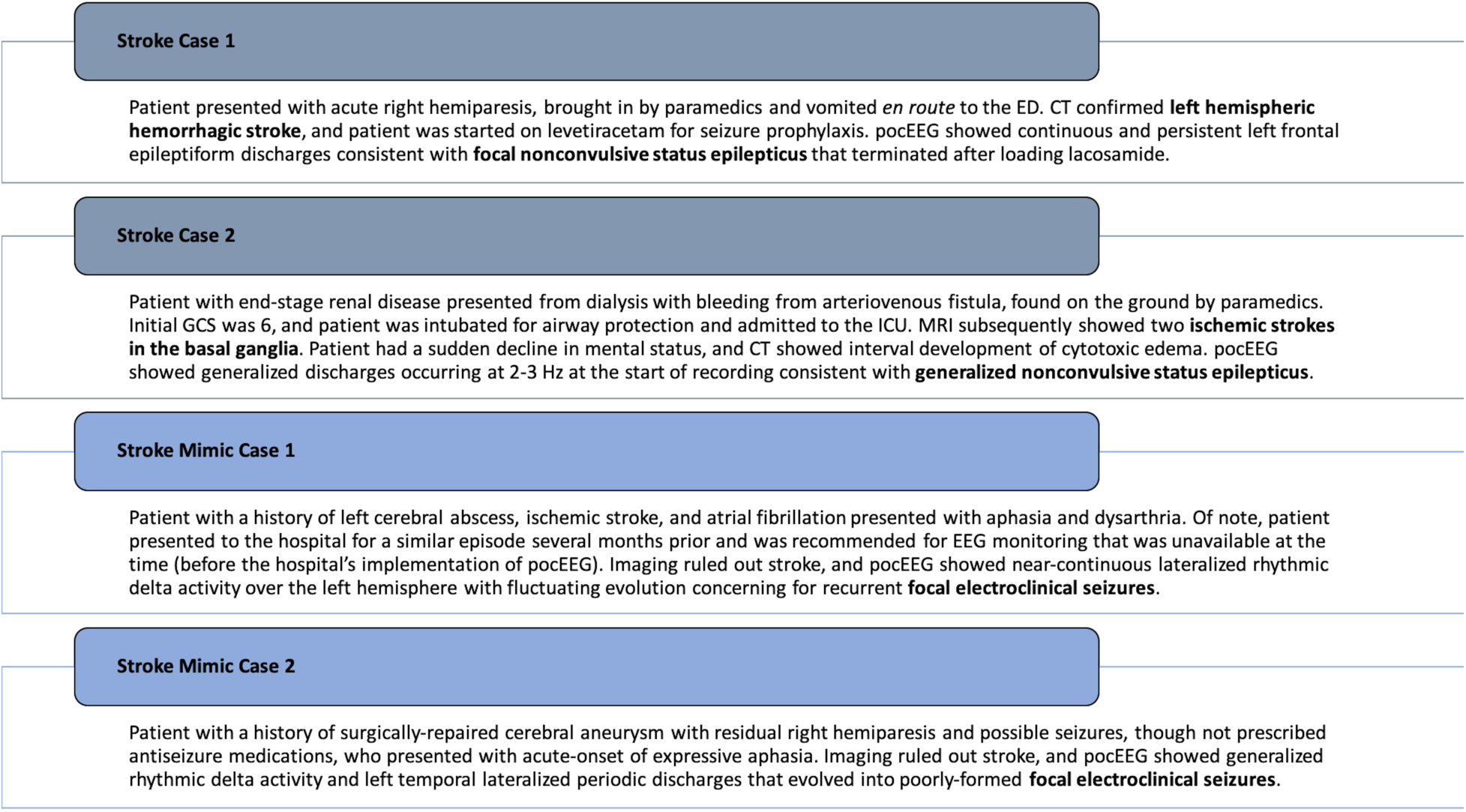
Clinical Vignettes of Patients with Ictal Point-of-Care EEG. Potentially identifying information (e.g., age and sex) were excluded from case descriptions. Abbreviations: CT, computed tomography; ED, emergency department; EEG, electroencephalography; GCS, Glasgow Coma Scale score; ICU, intensive care unit; MRI, magnetic resonance imaging.

## DISCUSSION

Using a combination of neuroimaging modalities, the stroke code is designed to detect acute ischemic or hemorrhagic strokes and identify candidates for reperfusion therapies to reduce stroke-related disability. However, neuroimaging alone is insufficient to either confirm acute symptomatic seizures and highly epileptiform patterns that might warrant treatment in patients with acute stroke or definitively diagnose seizures as a stroke mimic. Indeed, the bedside exam can also be limited when ictal and epileptiform activity do not produce overt clinical signs.^6^ Although EEG would be the ideal test for this indication, conventional EEG monitoring is often unavailable in emergency care settings, especially after-hours or in many community hospitals, and can be difficult to obtain in a timely fashion even in academic medical centers.^18^

In our community hospital, we employed a point-of-care EEG device with a reduced electrode array as an adjunct to acute stroke evaluation, enabling the rapid detection of ictal and highly epileptiform activity in 16% of patients with acute stroke and 34% of patients with stroke mimics. Similar to prior studies of stroke mimics^3,4,10^, seizure was the most common stroke mimic in our cohort, however its frequency in our cohort was noticeably higher, potentially due to the availability of rapid EEG monitoring.^38^ We also identified two patients with persistent expressive aphasia who were found to have focal seizures over the left (presumably language-dominant) hemisphere, consistent with reports of seizures with “negative” cortical signs mimicking acute ischemic stroke.^12^ Finally, point-of-care EEG monitoring was also able to rule out nonconvulsive seizures in the remaining patients without confirmed stroke on neuroimaging, many of whom would have been at increased seizure risk from toxic-metabolic and hypertensive encephalopathies or focal structural lesions.

While our focus was on describing the use of point-of-care EEG for rapid seizure detection in the stroke code context, it should be noted that EEG is also well-suited to guide the diagnosis of stroke. This is because characteristic electrographic changes – decreased fast (alpha) activity and increased slow (delta) activity – can be reliably detected within minutes of hypoperfusion/hypoxia and monitored using quantitative EEG as either an increased delta/alpha ratio or an increased brain symmetry index.^20,39–41^ These changes manifest on EEG even before ischemia is detected by either CT or MRI.^18^ To this end, there have been several efforts in recent years, such as the ELECTRA-STROKE trial^42^, to incorporate EEG devices into the prehospital and in-hospital evaluation to diagnose stroke, especially ELVO, and inform triage and treatment.^40,43–49^ Several of these studies even used reduced electrode arrays and dry electrode systems in lieu of the conventional EEG system (i.e., a full array of wet electrodes applied by a trained technologist) to circumvent the limitations of the current inpatient EEG infrastructure. While this is a promising direction for the field of clinical neurophysiology to expand its utility in cerebrovascular diseases, we feel that the established capability of EEG for detecting seizures and epileptiform activities and evaluating stroke mimics should remain as a core asset for electrodiagnostics in stroke.

### Limitations

This study is not without limitations. We employed a retrospective study design to identify a subgroup of patients from a larger study focused on point-of-care EEG monitoring in our community hospital, which limited the ability to obtain all desirable data, control for potential confounders, adjudicate diagnostic findings and final diagnoses, perform significance testing, and assess more long-term outcomes. We aimed to describe the standard of care at our center, in which point-of-care EEG is utilized at the discretion of the treating physician primarily to exclude nonconvulsive seizures and status epilepticus, but we could not enforce a standardized workflow for point-of-care EEG in acute stroke evaluation (e.g., timing relative to imaging and reperfusion treatments, reporting of stroke-related EEG changes, ASM treatment for particular findings). As such, this sample may represent a selected group of higher-risk patients more likely to have seizure-like presentations or clinical decompensation that prompted EEG monitoring. This is reflected in the relatively high frequency of stroke mimics diagnosed as seizures compared to prior studies of stroke mimic characteristics. Larger, prospective studies are needed to validate these findings, ideally with harmonized data reporting across multiple sites.

## CONCLUSIONS

Our findings support the utility of rapid EEG devices to screen for nonconvulsive seizures either as an acute sequela of stroke or as a stroke mimic.

## Data Availability

The data that support the findings of this study are available on reasonable request from the corresponding author.

## ACKNOWLEDGEMENTS

**CRediT authorship contribution statement:**

**Kapil Gururangan:** Conceptualization, Methodology, Investigation, Data curation, Formal analysis, Writing – original draft, Writing – review & editing, Visualization, Supervision. **Richard Kozak:** Conceptualization, Investigation, Data curation, Writing – review & editing, Project administration, Funding acquisition. **Parshaw J. Dorriz:** Investigation, Writing – review & editing.

**Declaration of competing interest:** Dr. Gururangan serves as a clinical and scientific advisor to Ceribell Inc. Dr. Kozak and Dr. Dorriz received financial support from Ceribell Inc. for this research.

**Sources of funding:** This project was supported by Ceribell Inc. The funders had no role in the design and conduct of the study; collection, management, analysis, and interpretation of the data; preparation, review, or approval of the manuscript; and decision to submit the manuscript for publication.

**Acknowledgements:** The authors would like to thank Matthew Kaplan, MD (Providence Mission Medical Center) and Josef Parvizi, MD, PhD (Stanford University) for their comments on the manuscript.

**Publication and Presentation History:** Findings from this report have been previously presented at annual meetings of the American Epilepsy Society (AES, 1-5 Dec 2023 in Orlando, FL), Society of Critical Care Medicine (SCCM, 21-23 Jan 2024 in Phoenix, AZ), and American Heart Association (AHA International Stroke Conference, 7-9 Feb 2024 in Phoenix, AZ).

**Data Sharing Statement:** The data that support the findings of this study are available on reasonable request from the corresponding author.

